# N2G calibrator: a cross-subject domain adversarial training framework for gait tracking from neural signals in Parkinson’s disease

**DOI:** 10.1101/2025.10.08.25337508

**Authors:** Jin Woo Choi, Helen M. Bronte-Stewart

## Abstract

Adaptive deep brain stimulation has enabled machine learning models to track motor states from neural signals with improved accuracy, aiming to provide electrical stimulation accordingly. Such data-driven techniques necessitate extensive user-specific data collection involving repetitive tasks and additional sensors to quantify continuous movements, due to variations in neural signals between individuals. In this study, we introduce Neural-to-Gait Calibrator, a cross-subject deep learning framework that leverages collective neural data to track gait performance of users with Parkinson’s disease. Our framework utilizes domain adversarial learning to calibrate target user’s neural signals using data from other individuals, removing the need for synchronous gait recording systems thereby enabling personalized model calibration outside equipped clinical settings. The framework’s effectiveness was demonstrated through a significant reduction in error rates compared to models trained with data from other individuals without calibration, achieving performance comparable to that of models trained directly with labeled target data.

## 1 Introduction

Adaptive deep brain stimulation (DBS) has recently gained attention as an emerging treatment for Parkinson’s disease (PD) due to its ability to sense neural signals in real time while simultaneously providing electrical stimulation to the brain through implanted leads [1–3]. Unlike continuous DBS which provides a consistent amount of stimulation throughout therapy, adaptive DBS adjusts stimulation based on predicted symptoms as reflected by neural signals [4]. By tailoring stimulation to effectively handle motor symptoms, adaptive DBS tends to reduce unnecessary stimulation thereby lessening potential side effects such as speech problems [5–7].

One of the actively researched symptoms for adaptive DBS is gait [8, 9]. Previous studies have addressed that local field potentials (LFPs) retrieved from the subthalamic nucleus (STN) contain information related to gait symptoms in people with PD (PWP). With signals in the beta frequency range (13-30 Hz) being the key neural features reflecting gait-related symptoms such as freezing of gait (FOG) and bradykinesia [10, 11], current algorithms driving the adaptive DBS utilize oscillatory neural characteristics to predict symptom severity and adjust stimulation accordingly. On many instances, beta power and beta bursts have been used with single- or dual-threshold algorithms to increase or decrease stimulation levels [12–14]. Due to the variations of such neural characteristics between individuals, many of the parameters and thresholds related to these neural features are determined manually by experts through data visualizations [15, 16]. As such approaches include assessing the movement performance of PWP and manually mapping it to visually identified neural signal characteristics, repeated trial-and-error testing is typically needed in order to effectively configure adaptive DBS therapy.

Recent literature has applied machine learning methods to provide more personalized therapy for individuals while also reducing the efforts required for setting up adaptive DBS [17]. These data-driven approaches tend to map neural signals to quantified kinematic traits using the data acquired from individuals, aiming to optimize parameters that had to be determined manually. Prior studies have focused on developing data-driven techniques related to gait. For instance, in our recent study, a deep learning-based regression model N2GNet was proposed to translate neural signals into gait performance that was quantified by measuring weight shifts during repetitive stepping in place tasks performed on force plates [18]. Deep learning has also been employed to correlate neural signals with activation of leg muscles, aiming to identify dysfunctional gait during walking [19]. Along with other methods linking neural signals to various movement symptoms [3, 20–22], machine learning approaches have shown their potential to provide a broader range of relevant neural inputs for adaptive DBS.

Although data-driven approaches have shown feasibility toward adaptive DBS therapy, these algorithms require considerable amounts of synchronized neural and kinematic data acquired specifically from the current user before they can be effectively employed [18, 19, 23, 24]. Particularly for algorithms involving neural signals, it is challenging to utilize previously collected data from other individuals to a new target user due to the variability in neural characteristics across individuals [25,26]. Furthermore, the data collection process requires extra sensors or measurement systems such as accelerometers, electromyogram sensors, dynamometric chairs, or force plates, to record kinematic data synchronously with neural signals [18, 19, 27, 28]. These requirements may hinder individuals from effectively using machine learning methods to establish or update their personalized adaptive DBS therapy, especially in urgent scenarios or at-home settings where only the implanted adaptive DBS system is available, with no supplementary sensors to simultaneously capture kinematic traits.

In this study, we present Neural-to-Gait Calibrator Framework (N2GCF), a cross-subject deep learning approach based on domain adversarial training that selectively aligns neural and gait data from source individuals with a target user’s neural signals for personalized gait tracking. Our framework does not require any gait measurements to be synchronously recorded with the neural signals from the target user, allowing them to establish a personalized gait tracking deep learning model simply by simulating walking in place. By utilizing a gradient reversal layer (GRL), adversarial training encourages the features extracted by the model not to be discriminative of whether they originate from the source or target domain, directing the model to learn domain-invariant features [29]. N2GCF adds our recurrent selective source scheme that utilizes maximum mean discrepancy (MMD) to continuously track and guide the framework to rely more on sources whose extracted neural features closely resembled those of the target individual throughout its training process. As the baseline model for our framework, we utilized one of the deep learning models introduced from the previous study which mapped STN LFPs to gait performance using simultaneously recorded neural data and quantified weight shifts collected during repeated stepping-in-place tasks [18]. We evaluated our framework by training it under two different conditions: an unsupervised approach where neural signals and quantified gait performance labels from source participants are used while only neural signals from the target participant are used to calibrate and finalize the regression model, and a semi-supervised approach where gait performance labels from the target participant are involved only in model selection for its final establishment. By evaluating the performance of our framework under both the unsupervised condition, where labels from the target are not considered at any phase of model establishment, and the semi-supervised condition, where target labels are utilized solely for selecting the best-performing model across epochs, we aimed to evaluate the framework’s practical applicability in reallife scenarios and quantify its capability in reducing feature discrepancies independently of the strategies used for model selection.

## 2 Results

### 2.1 Unsupervised condition results resembling real-life usage

The framework was trained and evaluated using the unsupervised condition, which resembles its real-life application where calibration occurs without requiring additional sensors for quantifying gait performance from the target user. In this condition, only the unlabeled neural signal data from the target participant were used throughout model calibration and selection, along with the labeled data from the source participants. As labels from the target are assumed to be unavailable for the unsupervised condition, our MMD-based model selection was used to select the final model for each target from those established throughout the training epoch (Figure 1 a).

**Figure 1:**
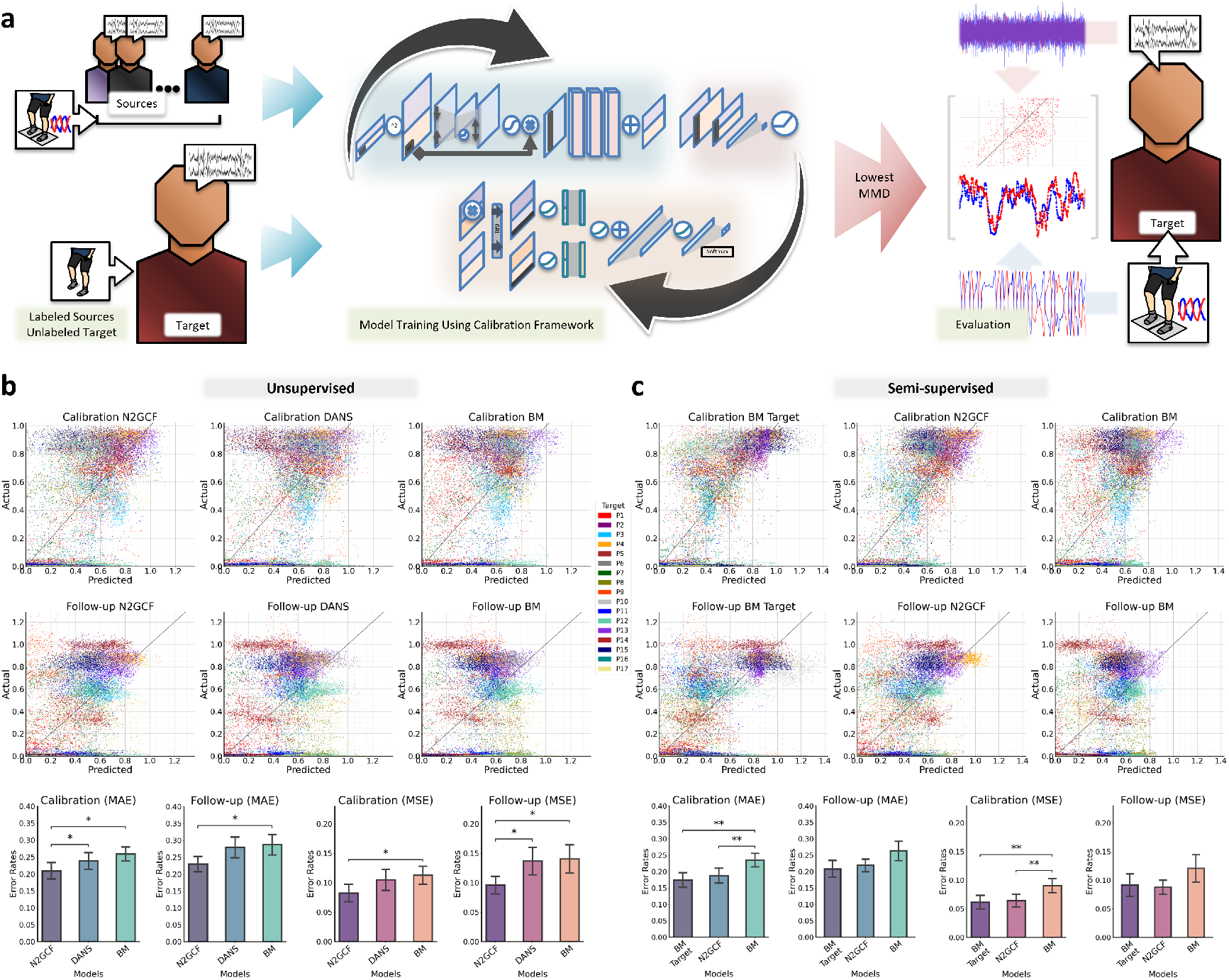
N2G calibrator training and evaluation. a Procedures for training and evaluating the N2G calibrator in the unsupervised condition using neural signal data and gait performance labels from source participants, calibrating with neural signal data without labels from the target participant. b Performance of N2GCF in the unsupervised condition, showing distributions of prediction results using the baseline model trained with labeled data from the sources without calibration (BM), using our domain adversarial training framework without our MMD-based recurrent source selection (DANS), and using our N2G calibrator framework (N2GCF). The x-axes for the spread indicate prediction results from the utilized models, and the y-axes indicate actual gait performance values derived from the force plates. The colors of the prediction samples represent results from the corresponding participant. Error rates for the predictions are quantified using mean absolute error (MAE) and mean squared error (MSE). c Performance of N2GCF in the semi-supervised condition, with distributions and quantified error rates of prediction results using the baseline model trained with labeled data from sources and without calibration from the target participant (BM), N2GCF, and the baseline model trained in a subject-dependent manner using labeled data from the target participant (BM Target) as opposed to using data from source participants. Error bars represent the standard error. ∗ *p <* 0.05, ∗∗ *p <* 0.01.

The distributions of prediction results from seventeen participants were explored for both calibration and follow-up datasets (Figure 1 b), where the evaluation of follow-up dataset represents the performance of the model after a substantial amount of time has elapsed since it was calibrated with the calibration dataset using the framework. Figure 1 b also shows the prediction results of our complete N2G calibrator framework, quantified using mean absolute error (MAE) and mean squared error (MSE) rates, compared to two other designs: our domain adversarial learning approach with no selection (DANS) that consistently considers all sources throughout training, and a baseline model (BM) that did not perform calibration with the target. As can be seen in the results, N2GCF obtained the lowest error rates from both calibration and follow-up sets, exhibiting a mean±standard error of 0.209 ± 0.024 MAE and 0.082 ± 0.015 MSE for the calibration sets, and 0.23 ± 0.022 MAE and 0.096 ± 0.015 MSE for the follow-up sets. Comparisons between

N2GCF and other approaches showed that our N2G calibrator was able to calibrate with significantly lower error rates than both the domain adversarial learning approach with no selection and the baseline model without adversarial learning in terms of MAE (DANS: 0.239 ± 0.024, *p* −*adj* = 0.02, *r* = 0.6315 and BM: 0.259 ± 0.021, *p*− *adj* = 0.0115, *r* = 0.6659). N2GCF also exhibited lower MSE rates than the other two, with statistical significance when compared to the baseline model (DANS: 0.105 ± 0.018, *p* −*adj* = 0.0697, *r* = 0.5396 and BM: 0.113 ± 0.015, *p* −*adj* = 0.0237, *r* = 0.62). For the follow-up data, the error rates were lower for the N2GCF compared to both DANS (0.279 ± 0.031, *p* −*adj* = 0.0605, *r* = 0.5511 for MAE and 0.137 ± 0.023, *p* −*adj* = 0.0386, *r* = 0.5855 for MSE) and BM (0.287 ± 0.03, *p*− *adj* = 0.045, *r* = 0.5741 for MAE and 0.141 ± 0.024, *p*− *adj* = 0.033, *r* = 0.597 for MSE). Statistical significance was observed between the three compared models with calibration data in both MAE and MSE (*p* = 0.0085 for MAE and *p* = 0.0194 for MSE, Friedman test), as well as in both with follow-up data (*p* = 0.0194 for MAE and *p* = 0.0096 for MSE, Friedman test).

### 2.2 Semi-supervised condition results reflecting the framework’s calibration capability

The spreads of prediction results were additionally explored using the semi-supervised condition to assess the potential calibration performance of our framework, independent of model selection approaches (Figure 1 c). The prediction performance of our N2GCF was explored along with the baseline model trained using source participant’s data without calibration, as well as with the baseline model trained using labeled data from the target participant (BM Target). In this semi-supervised condition where the target’s calibration data labels were made available for model selection, the model with the lowest L1 loss using the calibration data was selected for each target. As can be seen in Figure 1 c which quantified the error rates, performance comparisons with the three models showed that both BM Target (0.174 ± 0.022 MAE and 0.062 ± 0.012 MSE) and N2GCF (0.188 ± 0.023 MAE and 0.064 ± 0.011 MSE) had significantly lower error rates than BM (0.235 ± 0.021 MAE and 0.091 ± 0.012 MSE) with the calibration set (MAE: BM Target vs. BM with *p* − *adj* = 0.004, *r* = 0.7233 and N2GCF vs. BM with *p* − *adj* = 0.0025, *r* = 0.7463, MSE: BM Target vs. BM with *p* − *adj* = 0.005, *r* = 0.7118 and N2GCF vs. BM with *p* − *adj* = 0.0015, *r* = 0.7692). BM Target had lower error rates than N2GCF for the calibration set, but without statistical significance (MAE *p* − *adj* = 0.79, *r* = 0.2755 and MSE *p* −*adj* = 1.0, *r* = 0.2296). Additional evaluation using the follow-up set showed that BM Target (0.208 ± 0.026 MAE and 0.092 ± 0.019 MSE) and N2GCF (0.219 ± 0.019 MAE and 0.088 ± 0.012 MSE) had lower error rates compared to BM (0.263 ± 0.03 MAE and 0.121 ± 0.024 MSE) but without significant difference (MAE: BM Target vs. BM with *p* −*adj* = 0.1343, *r* = 0.4822 and N2GCF vs. BM with *p* −*adj* = 0.1044, *r* = 0.5052, MSE: BM Target vs. BM with *p* −*adj* = 1.0, *r* = 0.2296 and N2GCF vs. BM with *p adj* = 0.266, *r* = 0.4133). BM Target had a lower MAE than N2GCF without significance (*p* −*adj* = 0.6741, *r* = 0.2985), whereas N2GCF exhibited lower MSE than BM Target without significance (*p* −*adj* = 0.7305, *r* = 0.287). Statistical significance was observed between the three compared models with calibration data for both MAE and MSE (*p* = 0.0042 for MAE and *p* = 0.0033 for MSE, Friedman test), but was not observed with follow-up data (*p* = 0.056 for MAE and *p* = 0.1615 for MSE, Friedman test).

### 2.3 Comparison of calibrator’s predictions and beta power in relation to gait performance

To explore the efficacy of our N2G calibrator, we used the Kendall coefficient to compare the correlation of its predictions with gait performance against that of beta power, which was computed from 2-second time windows corresponding to the gait measurement and is a standard biomarker for PD-related motor symptoms. Figure 2 a shows the correlation results from the unsupervised N2GCF, the semi-supervised N2GCF, and beta power from both the higher- and lower-correlating of the two implanted DBS leads per participant. The results from the calibration sets showed that N2GCF from both conditions exhibited greater average correlation coefficients than the higher-correlating beta power (0.42 and 0.456 for N2GCF from the unsupervised and semi-supervised conditions, respectively, compared to 0.362 for the higher-correlating beta power). The coefficients measured with the follow-up data also showed greater average coefficients for N2GCF than the higher-correlating beta power, with values of 0.337 and 0.371 for N2GCF from the unsupervised and semi-supervised conditions, respectively, compared to 0.297 from higher-correlating beta power. The average coefficients for lower-correlating beta power were 0.237 and 0.182 in the calibration and follow-up data, respectively.

**Figure 2:**
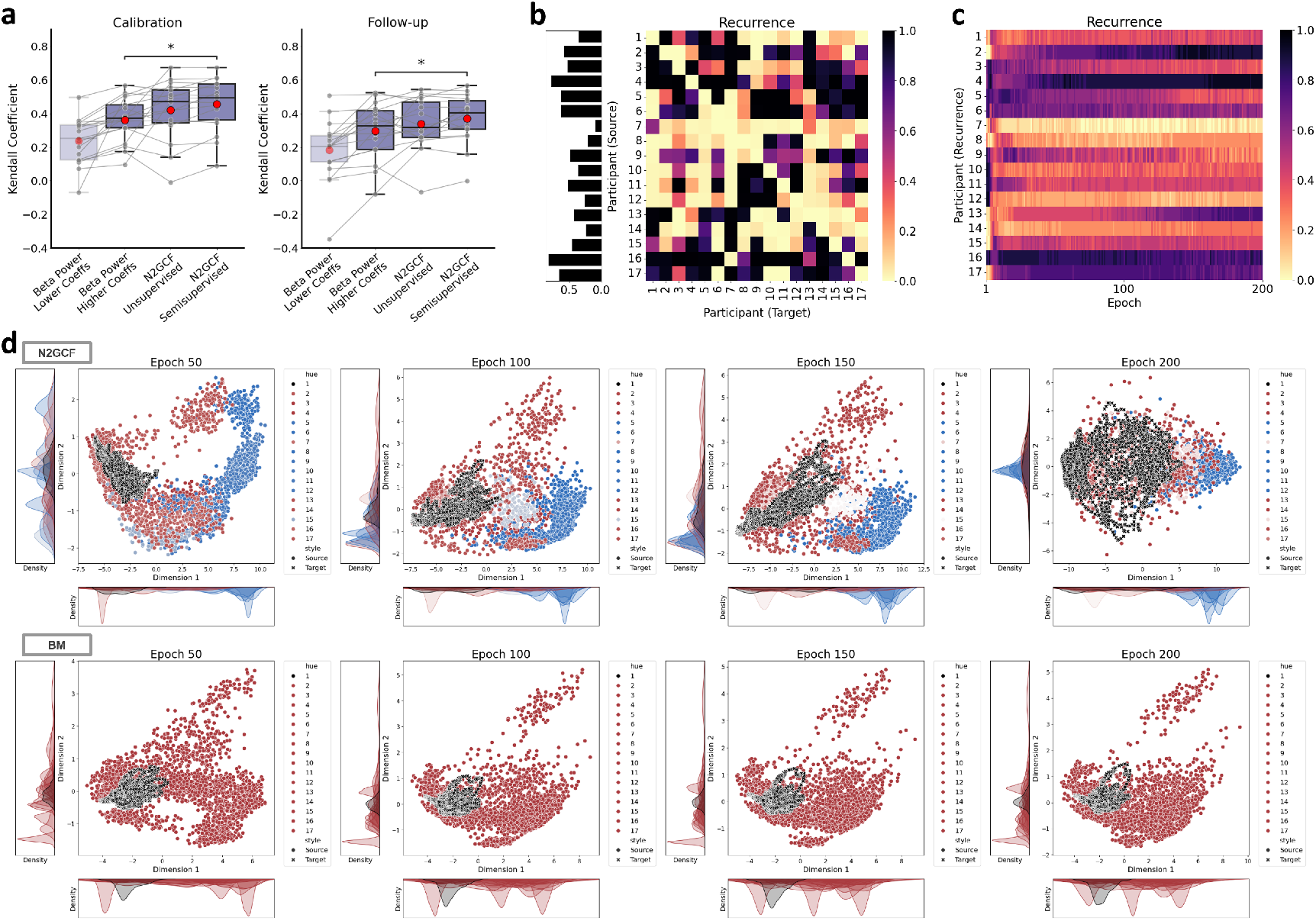
Results of Kendall correlation analysis and behavioral analysis for our N2GCF. a Comparisons of calibrator’s predictions and beta power with respect to their Kendall correlation with gait performance in the calibration and follow-up sets. b The recurrence ratio representing the frequency of each source being selected to be used for each target, until the epochs that achieved the minimum L1 loss with the calibration data. The x-axis represents the participant designated as the target, while the y-axis represents the participants as sources. The bar plot on the left shows the ratio of each source being used in total. c Average recurrence ratio of each source across targets on each epoch. The x-axis indicates the index of the epoch and the y-axis represents the participants as sources. d Feature distribution alignment between sources and target across training epochs for our N2GCF and the baseline model (BM). Principal component analysis (PCA) was applied to reduce dimensions for data visualization. For N2GCF, the color of data points from sources indicates their selection frequency up to the corresponding epoch, ranging from blue representing less frequent selection to red representing more frequent selection. BM utilizes all source participant data consistently in every epoch, as indicated in red across all sources. ∗ *p <* 0.05.

Statistical comparisons between the higher-correlating beta power, N2GCF from the unsupervised condition, and N2GCF from the semi-supervised condition showed that coefficients from the N2GCF in the semi-supervised condition exhibited significance compared to those from the high-correlating beta power in both calibration and follow-up data (calibration data with *p* − *adj* = 0.033, *r* = 0.597 and follow-up data with *p* − *adj* = 0.0386, *r* = 0.5855). Significance was not observed with the other comparison pairs.

### 2.4 Behavioral analysis of recurrent source selection

The recurrence ratio reflecting the frequency of the corresponding source being selected for each target until the epochs that reached minimum L1 loss with calibration data is shown in Figure 2 b. As can be seen in Figure 2 b, data from some participants were used more frequently for model training than others while data from some participants were rarely used, and the recurrence rate of sources varied when different participant data served as target. Figure 2 c further shows the average recurrence rate of the corresponding source being selected on the specific epoch index across targets, until the maximum number of epoch 200 is reached. The figure demonstrates varying results in source selection throughout the training epochs, indicating that the recurrent source selection was continuously adapted along with updated feature extractor as the training progressed.

Figure 2 d and Supplementary Figure 1 demonstrate the changes in the distribution of source and target neural features extracted from the models as training progresses, with the dimensionality of the features reduced using principal component analysis (PCA) for visualization. As can be seen in the upper figures that employed our N2GCF, the distribution of features from the target data increasingly overlapped towards those from more frequently selected sources as the epochs progressed toward the maximum epoch limit, indicating a reduction in discrepancies between the features from these sources and the features from the target. On the other hand, the lower figures that employed the baseline model exhibited relatively stable distributions throughout epoch progression, implying maintained discrepancies between features from the sources and the target.

### 2.5 Comparative analysis of prediction performance using other designs and approaches

Six design options derived during our framework development were employed to explore the effect on prediction performance of both domain adversarial learning and the selective adjustments of source influence throughout training. To evaluate the performance of these design choices independently of model selection methods, we acquired the performance of each design choice through the semi-supervised condition (Figure 3 a). Along with these design options, five domain adaptation approaches from computer vision and neural signal processing were adapted to our prediction task for benchmark comparisons.

**Figure 3:**
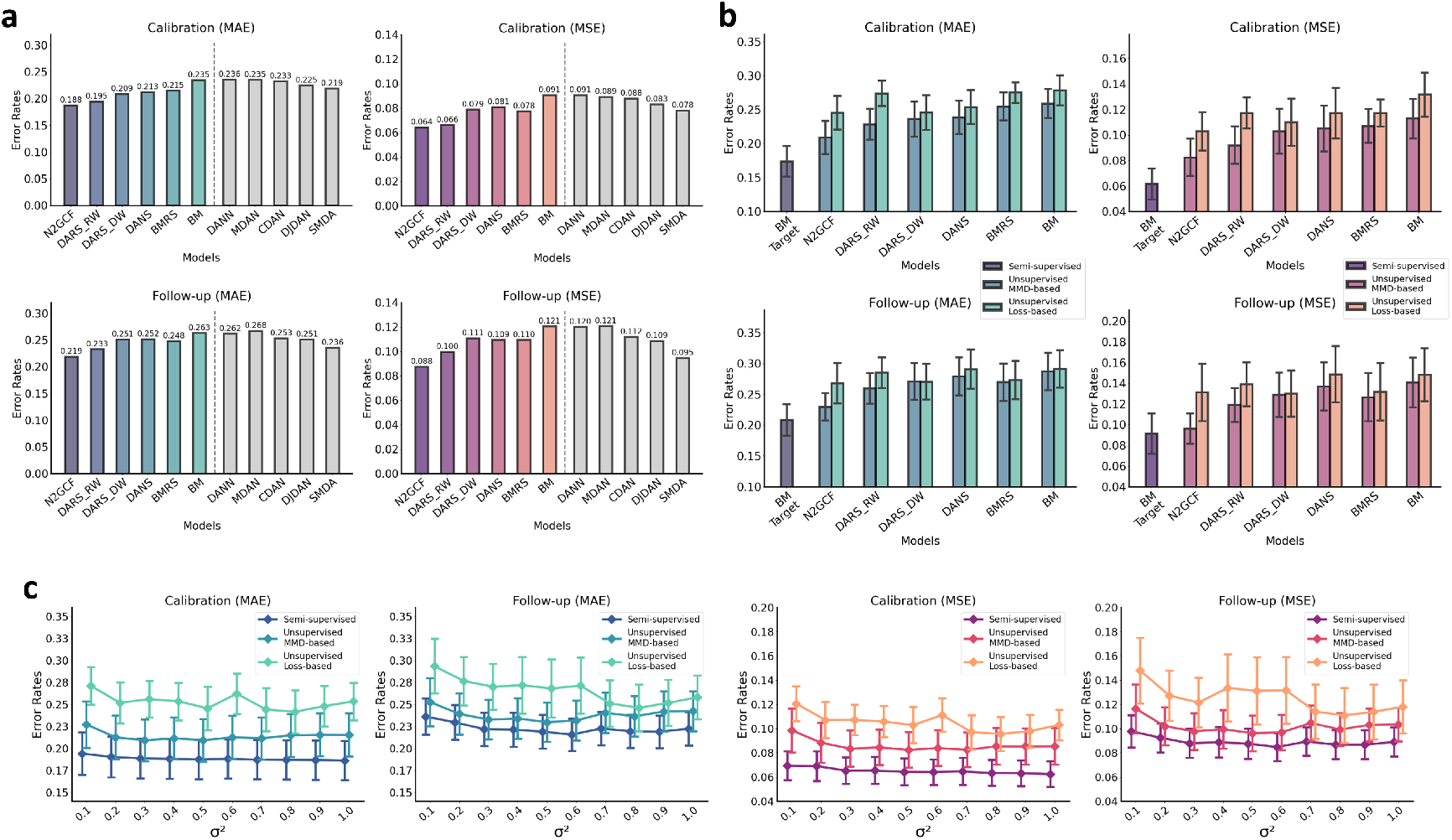
Performance analysis of different framework designs and conditions. a Averaged mean absolute error (MAE) and mean squared error (MSE) rates on the calibration and follow-up sets from the semi-supervised condition, including six different design choices related to the N2G calibrator framework and five different benchmark domain adaptation approaches. The design choices derived from our N2G calibrator differed in their use of adversarial learning techniques, and toward which backpropagation routes the recurrent source selection was applied to. b Comparisons between validation loss-based model selection and our MMD-based model selection in the unsupervised condition, using the framework designs derived from our N2G calibrator. The performance results from the baseline model trained with labeled target participant data in a semi-supervised condition are also provided as a reference. c Sensitivity analysis regarding changes in kernel bandwidth for the MMD-based measures. Error bars represent the standard error.

The results of comparing six different designs related to our framework indicated that our final N2G calibrator was able to outperform all other design choices on both calibration and follow-up sets in terms of both averaged MAE and MSE metrics. Specifically, average MAE and MSE error rates were improved from both the calibration and follow-up datasets when our recurrent source selection was applied towards backpropagation passing the regressor and feature extractor (BMRS vs. BM and DARS RW vs. DANS), whereas applying recurrent source selection only on the backpropagation path from the domain discriminator to the feature extractor exhibited smaller error rate reductions (DARS DW vs. DANS). Methods that utilized adversarial learning techniques exhibited lower error rates in both calibration and follow-up sets compared to when adversarial learning was not employed, regardless of whether our recurrent source selection was applied towards the regressor and feature extractor (N2GCF vs. BMRS and DARS RW vs. BMRS) or not (DANS vs. BM). Comparisons with five other domain adaptation approaches adapted to our regression task indicated that our full N2G calibrator framework structure was able to achieve lower average error rates in both our calibration and follow-up sets, with both MAE and MSE metrics.

Within the designs derived from our framework, two different model selection metrics for obtaining the model from those formed across training epochs were investigated under the unsupervised condition: one approach using L1 loss computed with the validation data from the source participants, and another using our MMD-based approach. As can be seen in both MAE and MSE metrics using the calibration set from Figure 3 b, the utilization of MMD-based approach for model selection resulted in lower error rates compared to using validation loss calculated with the data from sources. Similar patterns were observed for most of the investigated design choices when further evaluated with the follow-up data, where the models obtained using the MMD-based approach exhibited lower error rates than the models obtained using validation loss. Out of all the framework designs trained and selected in a cross-subject manner, N2GCF with MMD-based model selection exhibited the lowest error rates, lessening the performance gap towards the model trained and selected in a subject-dependent way and using labels from the target’s calibration data for model selection (BM Target).

### 2.6 Sensitivity analysis with different MMD kernel bandwidths

Additionally to explore the influence of MMD parameters in our approach, a sensitivity analysis regarding different MMD kernel bandwidths was performed. The bandwidth of the respective kernel was applied in both the recurrent source selection as well as in the model selection where the MMD was used. As shown in Figure 3 c, N2GCF from the semi-supervised condition consistently exhibited lower error rates in both the calibration and follow-up sets compared to the calibrator models established using two different selection metrics from the unsupervised condition. The calibrator using the semi-supervised condition also exhibited the lowest variability in error rates across different kernel bandwidths. As for the two unsupervised conditions, MMD-based model selections consistently outperformed models selected using validation loss from sources while also demonstrating lower variability across different kernel bandwidths.

## 3 Discussion

The aim of our N2G calibrator is to leverage existing LFP and weight shift data collected from other individuals, thereby reducing the data collection effort required from the target user in establishing their personalized gait-tracking deep learning model for adaptive DBS. The framework achieves this by directing the model to learn domain-invariant features, producing a calibrated feature extractor and a regressor that form a user-specific model capable of translating the target individual’s LFP signals into gait performance in real time. The results from the unsupervised condition showed that our framework was able to effectively calibrate data from source individuals to the target user, significantly reducing both MAE and MSE error rates in neural-to-gait translation with large effect sizes compared to the model without calibration. The calibration results obtained from the semi-supervised condition also demonstrated significantly lower error rates with large effect sizes for our N2GCF compared to the model that did not involve calibration, narrowing the performance gap towards the model trained directly with labeled data from the target individual. Our calibrator thus facilitates: 1) reducing the amount of data needed from the target user by substituting labeled training data from the target user with labeled data from source individuals, and 2) eliminating the need for synchronously recorded gait measurements from the target user, as only their neural signals are necessary for calibration. Our study possesses novelty in that we apply adversarial training to the regression task involving the use of DBS systems, where continuous weight shift values serve as labels for the framework model while using LFPs from the leads as input data. Particularly, our adversarial training strategy handles variations in LFP features between individuals through our MMD-based recurrent source selection, which aims to focus on sources whose neural features are more aligned with those of the target. With our N2GCF, we aim to simplify the overall process of establishing a deep learning-based personalized neural-to-gait translation model for future adaptive DBS systems, making it less demanding for the target user while preserving its accuracy.

It has been widely acknowledged in fields focusing on neural signal classification that training models with labeled neural data from other individuals without the target user generally results in notably lower performance compared to training with labeled data directly from the target user [30–32]. Due to this decrease in the model’s performance, extensive efforts are often put on data collection specifically from the target user despite the availability of data similarly collected from other individuals. Our study exhibited similar results, showing higher error rates for models trained in a subject-independent manner without calibration compared to those trained in a subject-dependent manner with labeled target data. As demonstrated in Supplementary Figure 2 representing data distribution plots using power spectral density (PSD) with different dimensionality reduction techniques known for preserving global structures including multidimensional scaling (MDS), PCA, and uniform manifold approximation and projection (UMAP) [33–35], this drop in performance may be due to misalignments between the data from different participants, which is an indication that neural signals are unique across individuals and possess varying characteristics. Developing a model capable of aligning extracted features between source and target data may be even more important for our study, as our objective involves a regression task that requires higher-resolution outputs than classification tasks. With our ultimate goal being the minimization of efforts required from the target user while maintaining the model’s performance, we aimed to address this problem using an unsupervised domain adaptation approach that reduces the discrepancy between the extracted features of informative sources and the target. Although the results of our adversarial training framework did not outperform the performance of models trained directly with the labeled data from the target user, it significantly reduced error rates through calibration, lowering error rates in the semi-supervised condition to a level that was not significantly different from those obtained using models trained with target user’s labeled data.

Our study provides an instance where simply relying on all the available data throughout model training, without considering the variability of neural signal characteristics between individuals and their feature alignment with the target participant’s neural characteristics, may result in degradation of prediction performance. It may be possible that some sources are entirely uninformative for the target participant or may even be damaging to model training, leading to greater error in the target participant model’s performance. Several factors may contribute to the distinctiveness of an individual’s signals, including variations in aperiodic signal components influenced by an individual’s physiological state, causes due to neural plasticity, and differences in signal quality arising from lead placement and positioning [36–39]. As these factors unrelated to movement collectively form the intrinsic nature of neural signals and may introduce confusion into the model, our MMD-based recurrent source selection approach aimed to identify the sources whose extracted features could not align closely with the target based on the updated model from the latest epoch, and subsequently limit the influence of these sources in the following epoch to prevent the model from learning signals with irrelevant characteristics. The effect of our recurrent source selection could be demonstrated from the results of our semi-supervised condition which aimed to investigate the calibration performance of the framework itself independently of model selection methods. By further comparing with other possible framework design options that either had recurrent source selection only in terms of regression-related features, which is limiting certain sources from influencing the feature extractor and regressor, or only in terms of domain-related features, which is limiting certain sources from influencing the feature extractor via the domain discriminator, our model ablation study results indicated that not all sources necessarily contribute to performance improvement as they calibrate toward the target. Most importantly, using our N2G calibrator’s recurrent source selection along with adversarial learning noticeably enhanced calibration performance compared to when the adversarial learning strategy was not used and also outperformed the case where adversarial learning was used without any source selection, highlighting the effect of our overall approach for regression-based gait prediction that aims to generate continuous values. Apart from previous studies involving neural signals that applied adversarial training to classification tasks [40–42], our study emphasizes the importance of utilizing neural data from individuals whose patterns align more closely with those of the target participant and addresses its feasibility in the regression task.

It is important to note that our feature extractor, domain discriminator, and regressor architecture was incorporated into the approaches we used for benchmark comparisons. This adaptation was essential as many established domain adaptation methods, particularly within neural signal processing, were originally developed for classification rather than for our regression task of tracking gait performance represented with labels of continuous values. While the results of these adapted models should thus be interpreted carefully, our analysis conveyed two notable findings. As expected, approaches that enabled the domain discriminator or discriminators to consider conditional features derived from the feature extractor and the regressor, simultaneously along with marginal features, exhibited lower error rates than others. Unexpectedly, however, approaches that only considered marginal features outputted from the feature extractor did not improve, and in some cases, performed worse than the baseline model that did not involve any domain adaptation process. This shows that for regression tasks involving variable features across domains such as neural signals, solely focusing on marginal distribution alignment may not be effective for its domain adaptation.

The results of using our MMD-based model selection, an approach used in our framework to determine one of the models generated from training epochs for final establishment, show that selecting models based on validation loss from source data may not serve as an optimal strategy for regression tasks. With labels from the target data unavailable, unsupervised conditions require more complex metrics for selecting reliable models. While using validation loss is a common practice for classification tasks in deep learning and is also occasionally used in unsupervised domain adaptation with the data from sources [43–45], our comparison between validation loss-based selection and our MMD-based selection indicated consistently higher error rates for the validation loss-based approach across multiple model designs. Aligning with previous studies that highlight the importance of model selection metrics for unsupervised domain adaptation in classification tasks [46–48], our study emphasizes that using suitable metrics for obtaining final models is also critical for our regression task. Comparable to the validation loss-based selection approach that does not use additional complex methods, our current model selection directly utilizes features already available in our framework through our feature extractor and regressor, and simply selects the models with lowest average MMD calculated. By taking conditional features into account that consider the distribution of regression outcomes through the use of predicted outcomes from the regressor, which is absent in the validation loss-based model selection, our MMD-based approach exhibited lower error rates. Beyond our simple metric, further enhancements in model selections can be done in advance specifically focusing on our regression task, where the distribution of continuous label values is uneven across trial recordings due to the inconsistent elicitation of symptoms in PWP.

Compared to the error rates assessed using calibration data, results from the follow-up data exhibited relatively smaller performance differences between models with weaker statistical measures, especially in the semi-supervised condition. This may be due to the fact that neural signals within an individual can change over extended periods of time. It has been observed in prior works that the aperiodic components of neural signals in PWP change over time depending on multiple factors including severity of their symptoms, cognitive impairment, and influences from neural plasticity [37, 49, 50]. Changes in neural signals may negatively impact model performance, as can be seen from our results showing increased error rates for the follow-up data, which was recorded at least one month after the calibration data, compared to evaluations with the calibration data itself. Such performance degradation due to shifts in the domain space also implies that using data-driven models may require periodic adjustments to their parameters in real-world settings.

Along with the associated degradation in the deep learning model’s gait tracking performance shown in our study, these variations in an individual’s neural signals over time thus highlight the need for a practical recalibration method such as our N2G calibrator. Conventional subject-dependent deep learning methods require synchronous recordings of neural signals and gait metrics, a process that would necessitate clinic visits with specialized equipment. In contrast, our N2G calibrator is designed to shift this paradigm by allowing users to update their personalized gait tracking models independently of location, even from their own homes. When applied in a real world setting, our framework would instruct the user to simulate a simple stepping in place task to acquire only their neural signals, without any specialized equipment to measure gait at the same time. The calibrator would then utilize these neural recordings in conjunction with an existing database to create the personalized neural-to-gait translational model. As neural signals can change over time and may thus require periodic model recalibration after certain periods, our calibrator approach could allow users to update their models from anywhere they can perform walking in place, removing the burden of repeated clinic visits and making the technology more accessible.

Our analysis using Kendall correlation demonstrates that the N2G calibrator provides a more advanced biomarker for tracking gait than beta power, which is used in many studies to monitor motor symptoms in PD [10–12, 17]. The predictions from the calibrator established with the unsupervised condition exhibited a stronger average correlation with gait than beta power, and this improvement was even statistically significant for the calibrator established in the semi-supervised condition. A key distinction is that our calibrator quantifies gait performance, whereas beta power is limited to indicating relative changes in movement. This transition from a relative indicator to a quantitative metric suggests our framework can provide clinicians with more direct and comprehensive information for tracking disease progression or treatment efficacy. By using Kendall’s coefficient that evaluates the ordinal association rather than the magnitude of error, we could directly compare our quantitative predictions against the relative fluctuations of beta power. This analysis, which provides a measure of clinical relevance by benchmarking against beta power that is inaccessible through MAE and MSE, showed that our calibrator still had a stronger association with gait performance than beta power, even without considering magnitude. The coefficient results also imply that the calibrator training process was able to automatically identify and capture key neural characteristics, including those from beta fluctuations, from the broad frequency range of neural signals. With MAE and MSE results demonstrating its greater accuracy in predicting the magnitude of gait performance than other framework designs, and the Kendall’s coefficient analysis showing its stronger relative correlation with gait compared to beta power, our evaluation highlights our calibrator as an enhanced and clinically relevant biomarker for gait tracking with neural signals.

There are some limitations and potential areas for improvement in our N2G calibrator framework. One limitation in our study is the small number of sample sizes used for training and evaluating our framework. As our datasets include a total of seventeen participants, our statistical analyses may not be powerful due to sample size limitation, and thus the results may need careful interpretation. Collecting additional recordings from more participants may further improve the performance of our N2GCF by providing a wider range of neural characteristics and broadening selection choices for calibration with the target’s neural signals. Another possible limitation is our fixed approach to recurrent source selection, where we simply limited selections to exactly half of the available source participants in each and every epoch. A more detailed approach that allows for flexibility in adjusting the number of selected sources in each epoch, while also considering the feature distributions between sources, may lead to further improvements in prediction performance. Although our findings conveyed that using all available sources does not lead to better outcomes by employing our MMD-based recurrent source selection, there still remains room for further optimization of our selection metrics. If more data can be acquired from other participants, future works may additionally dedicate efforts to developing enhanced solutions for model selection within our framework. While the current study primarily focused on exploring the feasibility of our adversarial training strategy for calibrating neural signals between source and target individuals, model selection methods also play a crucial role for the framework’s practical applicability, as we have observed in our study results. As the current study has limited number of participants, additional approaches focusing primarily on the development of model selection should be carried out in advance with larger participant sample sizes, such that the differences in performance between various model selection approaches can be clearly evaluated and analyzed in detail. Lastly, improving the feasibility of our framework for DBS implants may also be held for its real-life applicability. A key limitation for the real-world application of the deep learning models in neural signals, including our framework, is their high computational cost that restricts their use in implantable devices. Using our current N2G calibrator as a benchmark, further work can focus on developing more lightweight frameworks in order for them to be embedded directly into future DBS systems.

In this study, we propose a deep learning-based framework that aims to map STN LFPs into gait performance, necessitating only unlabeled LFP data from the target participant by utilizing labeled data from other source participants. Our N2GCF employs adversarial training and recurrent source selection to track and identify sources whose neural signal features closely align with those of the target as training progresses, calibrating their neural signals toward the target. Our approach demonstrated its efficacy by enhancing prediction performance compared to those without calibration, and also showing better prediction performance than other alternative adversarial training approaches. Our method also eliminated the need for gait quantification systems with sensors, such as inertial measurement units (IMUs) or force plates, that had to be synchronously measured with neural signals. The use of unlabeled data from the target user, in contrast to using labeled data, thus simplified the calibration process for deploying deep learning models to adaptive DBS systems. Overall, our N2G calibrator contributes toward a simplified calibration procedure for deploying deep learning-based adaptive DBS systems, enabling users to update their personalized models wherever they can simulate a simple stepping in place task.

## 4 Methods

### 4.1 Participants

The data used for this study is a subset of the data from a previous study [18]. Data collected from seventeen participants diagnosed with PD, who were able to perform all three stepping in place (SIP) trials without exhibiting gait freezing throughout the entire duration of any trial, were included in the study. Participants were implanted with an investigative neurostimulator (Activa PC+S, Medtronic, PLC) that was connected to bilateral DBS leads placed in the STNs. The study was conducted in accordance with the Declaration of Helsinki. All participants gave written informed consent prior to participating in the study. The study was approved by the Food and Drug Administration with an Investigational Device Exemption (G130186) and by the Stanford University Institutional Review Board (25916 and 30880).

### 4.2 Data description

The data from each participant included a total of three SIP trials, where the trials were conducted in separate visits with at least a one month gap in between (mean interval: 5 months between the earliest and intermediate trials, 7 months between the intermediate and latest trials). Participants performed each SIP trial in an off-medication state and without DBS stimulation. Prior to the SIP trial, the stimulation was in OFF state for at least 15 minutes and the participants were asked to stand on the force plate system while wearing a safety harness.

A single SIP trial included one continuous period of repetitive stepping in place. In the initial phase of the SIP trial, participants were asked to stand still on the two force plates and maintain motionless state. Upon receiving a start cue, participants were instructed to begin stepping alternately in place on the force plates at their own comfortable pace (Figure 4 a). After repetitive stepping in place for approximately 100 seconds, participants received a stop cue informing them to cease movement and remain standing still on the force plates.

**Figure 4:**
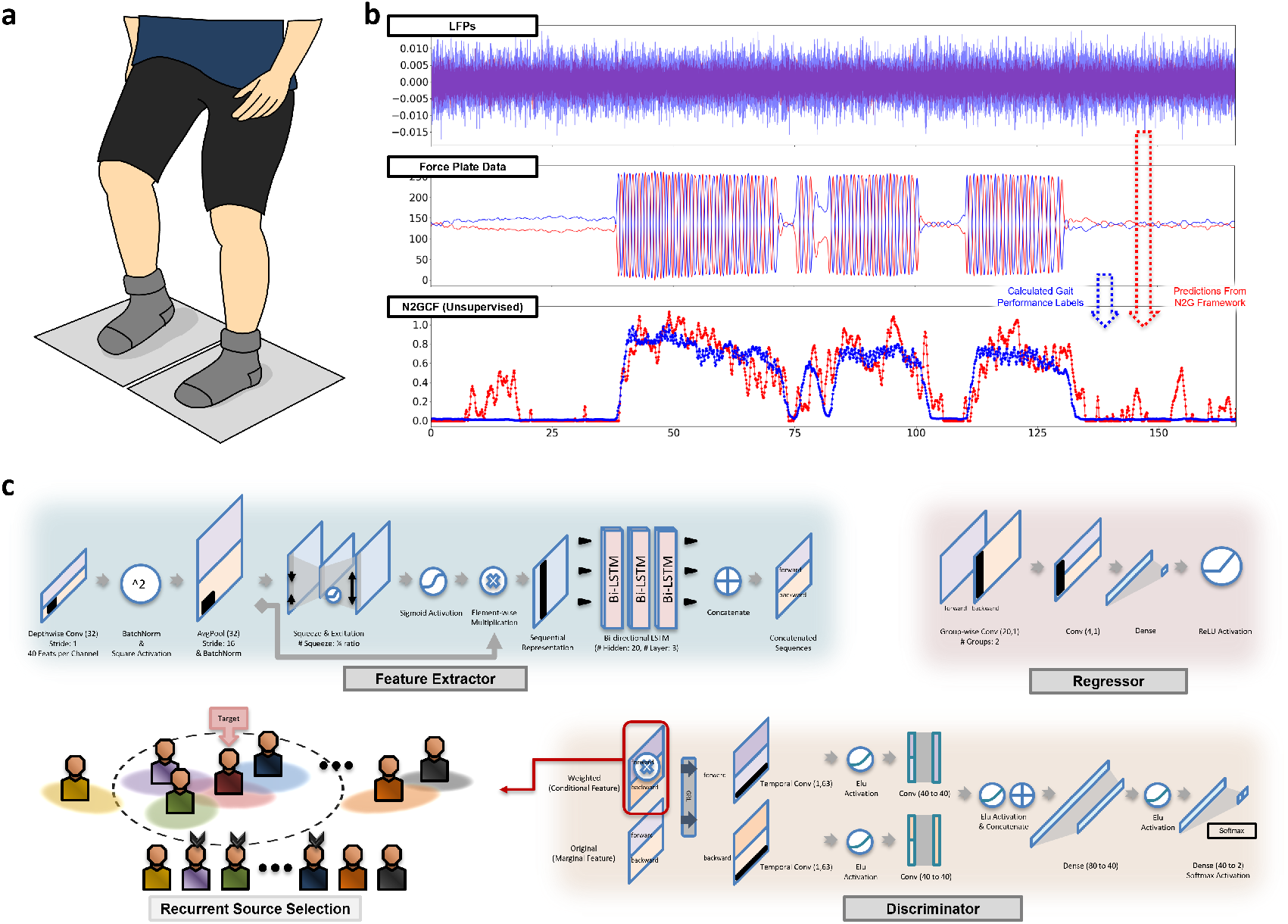
Illustration of the stepping in place trial and a description of the detailed components of our N2GCF. a Participants performed repetitive stepping in place during the trial, where they were instructed to raise their legs alternatively as if they were walking while staying on the two force plates. b Recorded LFP and force plate data from a single stepping in place trial. The force plate data were used to quantify gait performance and served as labels for the dataset, which was used for training and evaluating the framework. c The architecture of our N2G calibrator framework composed of feature extractor, regressor, and domain discriminator. Our MMD-based recurrent source selection was applied at each epoch to focus on training data with source participants whose weighted features more closely resembled those of the target participant.

Both LFPs from the two DBS leads and data from the force plates were recorded synchronously throughout the SIP trial, with the DBS stimulation maintaining OFF state until the end of the trial.

### 4.3 Dataset formation

The LFP signals retrieved from the participants were recorded with a 422 Hz sampling rate, and were downsampled to 211 Hz. A band-pass filter between 8 to 100 Hz frequency range was applied to the LFP signals to exclude frequency bands that may contain excessive noise, while still preserving a broad frequency range for the deep learning model to consider. The force plate data was acquired using either Neurocom (Neurocom Inc, Clackamas, OR, USA), which was measured with 100 Hz sampling rate, or Bertec (Bertec Corporation, Columbus, OH, USA), which was measured with 1000 Hz sampling rate. In cases where Bertec was used to measure ground reaction forces, the data were further downsampled to a 100 Hz sampling rate to match the sampling rate of the Neurocom data. The force plate data were low-pass filtered at 2 Hz to remove possible noises not involved with weight shifts associated with alternative stepping.

Gait performance was quantified using weight shift measurements, following methods similar to those described in the previous study [18]. The force plate data were first rescaled by dividing by the participant’s weight, estimated from the data retrieved while the participant was standing still on the two force plates. Data from the two force plates at each sampling time point were first integrated into one value by considering the greater one and then symmetrically reflecting values from one force plate around the value equivalent to half the weight of the participant. Quantified changes in the resulting sequence of values over a specific time window was used to represent participant’s weight shifts for the corresponding time interval. These procedures were done considering possible variations in participants’ weight across different visits, as well as limitations of the force plate in tracking weight shifts when one foot is entirely off the plate.

The dataset from each SIP trial was structured such that the last 5 seconds of the LFP signals at each time point served as the data, and the representative weight shift value of the last 2 seconds at the corresponding time point served as the label for each sample. Samples were generated at every 1 second stride time point when the participant belonged to the source, and samples were generated at every 0.1 second stride when the participant served as the target. This was done in order to balance the amount of source and target data, as data from multiple participants were included as sources whereas data from only one participant served as the target for model training. Hence, the data structure simulates tracking of gait performance using the most recent 5 seconds of neural data, with the output generated at 0.1-second intervals (Figure 4 b).

### 4.4 N2G calibrator framework architecture

Our overall N2GCF architecture extends one of the model designs introduced in the previous study [18] and can be separated into three different components each having a specific purpose in line with adversarial training from prior work [29]: a feature extractor, a regressor, and a domain discriminator (Figure 4 c). The feature extractor receives LFP signals from the two leads as input and learns relevant features based on the labeled LFP data from the sources and the unlabeled LFP from the target. The regressor takes the features from the feature extractor and maps them to gait performance values. The domain discriminator, which considers both the features from the feature extractor and the modified features processed using the output of the regressor, aims to learn domain-related features from the given inputs. The major role of the domain discriminator is to predict whether the given features originate either from the sources or the target. By including a GRL, the domain discriminator is used to derive the feature extractor to learn features that are invariant across domains. The detailed architecture of each component forming our framework is designed as follows:

- Feature extractor: For the feature extractor, we employed a design similar to one of the models in a previous study [18], which also aimed to predict gait performance from STN LFP signals but in a subject-dependent manner. The feature extractor used for our study begins with a feature extraction block composed of a depthwise one-dimensional convolutional neural network (CNN), a batch normalization layer, a square activation function, an average pooling layer, and another batch normalization layer. This feature extraction block is followed by an oscillatory feature squeeze-and-excitation block, where the feature dimension is squeezed and excited for each temporal dimension sample using shared encoding and decoding layers, with ReLU and Sigmoid activations placed in between and after these layers, respectively, maintaining its output shape the same as its input shape. The feature extractor lastly includes a bi-directional long short-term memory (LSTM) with three stacked layers, which inputs rescaled features derived by performing element-wise multiplication between the outputs obtained before and after the squeeze-and-excitation block. The resulting outcomes after this bi-directional LSTM block serve as the finalized features from the feature extractor, which are then used for our regressor and domain discriminator.
- Regressor: The regressor includes a groupwise CNN that separately extracts forward and backward LSTM features, followed by another CNN layer that merges forward and backward features, a dense layer, and a ReLU activation function. Using the output features from the feature extractor as input, the regressor produces a single-value outcome per data sample that represents predicted gait performance.
- Domain discriminator: Our domain discriminator consists of a GRL, followed by convolutional and dense layers with Elu activation functions placed between each of these layers, and a Softmax activation. The features from the feature extractor are first multiplied with the output value from the regressor, representing conditional features for the domain discriminator. Along with the features prior to multiplication that represent marginal features, these two types of features are first separated and concatenated into those originated from either the forward direction or backward direction of the LSTM. With the temporal dimension of the feature as the width and the other remaining feature dimension as the height of the input plane, the features from each LSTM direction were separately processed with their unique set of two consecutive CNN layers. In the first layer, features across the width dimension are compressed into a single feature value, whereas the second layer generates features for the corresponding LSTM direction by combining the compressed feature values from conditional and marginal features. Specifically, the aforementioned two CNN layers apply strides and kernels spanning the entire width and height dimensions, respectively, such that the layers behave similarly to dense layers applied across the corresponding dimensions. The resulting features from the forward and backward directions of the LSTM are finally concatenated, and the two dense layers are lastly used to complete the classification task.

For the framework in this study, bias was not used in the layers of the feature extractor except for its squeeze-and-excitation block, and it was also not used in the layers in the regressor. As a whole, the framework architecture takes LFP signals of a 5-second time window from the two leads through the feature extractor and produces a single gait performance value corresponding to the last 2 seconds of the time window from the regressor (Figure 5 a), while information regarding the discriminability of features related to the source or target is made available by the domain discriminator for adversarial training.

**Figure 5:**
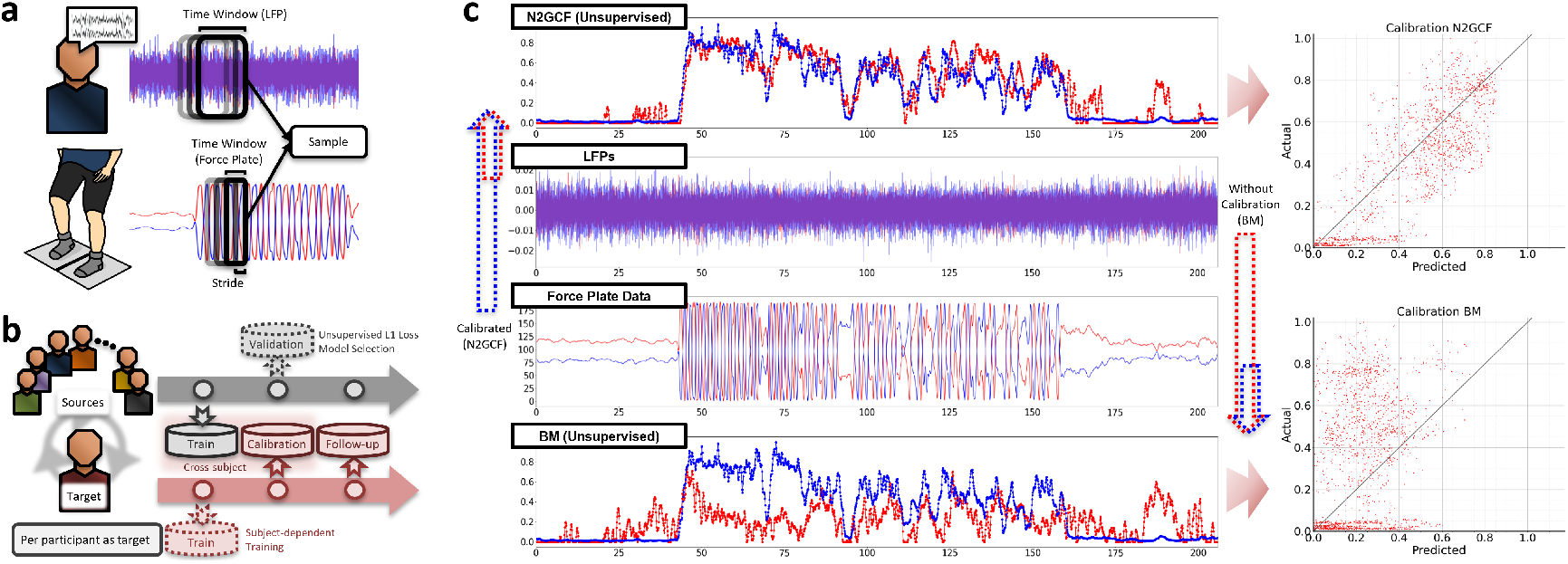
Description of datasets. a Data recorded from each trial was formed as a dataset consisting of samples retrieved throughout each stride time point, where a sample includes neural signal data and labels representing quantified gait shifts of a specific time window. b Distribution of datasets for training the framework, where data from the first trials of the source participants served as a training dataset, while data from the second and third trials of the target participant served as calibration and follow-up datasets, respectively. The first trial data from the target participant was used for training when the model was intended to be trained in a subject-dependent manner, and the second trial data from the source participants was used when the model selection was held with validation loss in the unsupervised condition. c Retrieval and comparison of performance results on the explored framework designs after the models were established. The data from the calibration and follow-up sets were inputted to the trained framework, and the results were quantified into error rates using mean absolute error (MAE) and mean squared error (MSE).

### 4.5 Adversarial learning strategy

We denote the data from participants used as sources by 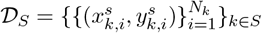, where *k* represents each individual participant within a set of source participants *S, N*_*k*_ is the number of data samples from participant *k*, and 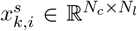a nd 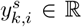 are the LFP data and gait performance label of a sample *i*, respectively, with *N*_*c*_ and *N*_*l*_ denoting the number of LFP channels and the temporal length of the LFP signals, respectively. We define the data from the unlabeled target domain as 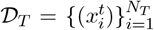 with 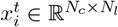, where *N*_*T*_ represents the number of data samples from a target participant *T*.

The aim of the feature extractor and the regressor for our N2GCF is to minimize the regression error using data from the selected source participants. As we approach our model training in an unsupervised manner, the regressor utilizes L1 loss with the source data:

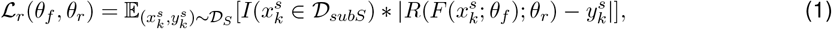

where *F* and *R* are the feature extractor and regressor, respectively, 𝒟_*subS*_ is the data from source participants *subS* ⊂ *S* that were selected in the previous epoch based on our maximum mean discrepancy (MMD)-based recurrent source selection, and 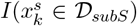 is an indicator of either 1 if 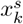 is from the subdomain 𝒟_*subS*_ or 0 otherwise. Thus, the loss was calculated using data from the subset of source participants whose extracted features had relatively smaller discrepancies with the target participant compared to those not included in the subset.

The purpose of the domain discriminator is to identify features that distinguish whether the data originates from sources or target. Our domain discriminator takes in not only the features directly obtained from the feature extractor but also the features adjusted with the outcomes from the regressor, representing marginal and conditional features for the data distribution, respectively. The inputs for the domain discriminator are generated by concatenating the two feature types:

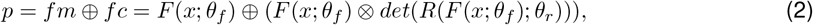

where *fm* and *fc* are marginal and conditional features, respectively, ⊗represents the concatenation of the two features maintaining the shared temporal dimension, ⊗indicates a product operator, and *det* indicates a detach operator that prevents its gradients from being backpropagated, treating its inputs as constants. By considering both the marginal features, which represents the discrepancy between extracted features, and the conditional features, which further emphasize the distributions of features conditioned along with the outputs from the regression, our domain discriminator aims to align both the distributions of the learned features and their associations with the outcome results, which have been shown to be effective approach for domain adaptation in previous studies [51–54]. The domain discriminator takes the resulting concatenated features as input and is trained with loss aimed at performing binary classification on each sample:

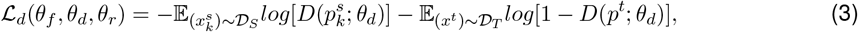

where *D* represents the domain discriminator, and 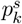 and *p*^*t*^ are the concatenated features derived from the source data 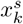 and the target data *x*^*t*^, respectively. Our domain discriminator thus learns to discriminate whether the input belongs to the source or target domain, using all available source data regardless of the MMD-based recurrent source selection results.

Through our adversarial learning approach, we aim to have the feature extractor to prevent itself from learning domain-related features exclusively from the sources that were selected through our recurrent source selection. With the use of a GRL, the adversarial loss applied to the feature extractor is noted as follows:

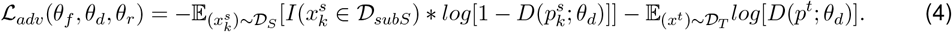

Thus, the overall optimization procedures for our N2G calibrator is denoted as:

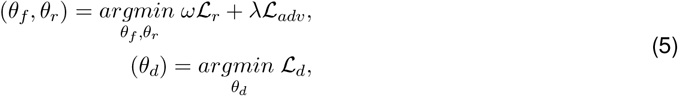

where *ω* is a re-weighting constant for the loss from the regression, measured as |*S*| */* |*subS*|, and *λ* is a parameter representing the weight of influence of the domain discriminator on the feature extractor. To take into account unstable feature extraction in the early framework training phases, we followed a method to adjust the parameter *λ* based on a previous study [29]:

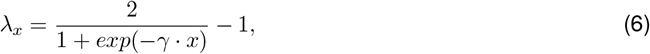

where *γ* is a fixed constant of 10, and *x* is the ratio of the current batch relative to the total number of batches scheduled, considering maximum epoch limit. Thus, the parameter gradually increased as the training progressed toward maximum epoch.

### 4.6 Maximum mean discrepancy-based recurrent source selection

Our recurrent source selection scheme is designed to guide our N2GCF to focus more on source participants whose neural characteristics are more similar to those of the target participant throughout the training process. As the framework progresses through adversarial training, we utilize MMD [55, 56] computed from the outcomes of the feature extractor and regressor to measure discrepancies between sources and target features, serving as an indicator of which source data should be considered further. Source selections occur after each epoch using training and calibration data of source and target participants, respectively, and the updated subset of sources is taken into account for training the framework in the subsequent epoch. All sources are included in the very first epoch of training, as the subset of sources has not yet been established.

We recall *fc* from the definition of our adversarial learning strategy, where 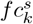 indicates conditional features from source participant *k* ∈ *S*, and *fc*^*t*^ represents conditional features from the target participant. Let 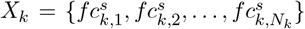 be the set of conditional features from source participant *k* and 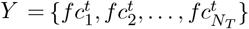 be the set of conditional features from the target participant. Here, we calculate the MMD between the two distributions *X*_*k*_ and *Y*, measuring the distance between these two feature sets from a reproducing kernel Hilbert space (RKHS) [57].

Let *κ*(*x, y*) be a positive definite kernel function for the Hilbert space, which calculates the similarity between *x* and *y*. The MMD between our two sets *X*_*k*_ and *Y* can be estimated as:

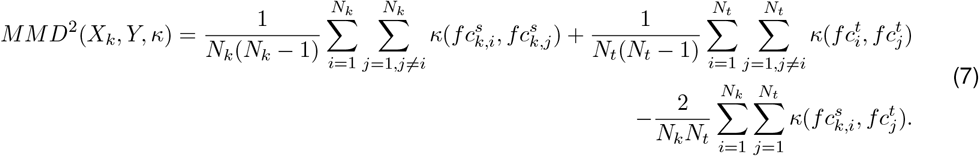

We utilized a Gaussian kernel for our study, which represents data into an infinite-dimensional space and is thus widely applied in the field of domain adaptation that prefers high-dimensional feature representations [58–60]:

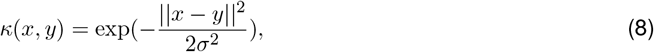

where *σ*^2^ was set to 0.5 for our study. By using the MMD measures between each source participant and the target data as the criterion for selection, we compose a subset of source participants that would be considered for our framework model training in the subsequent epoch:

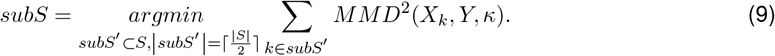

Inspired by the previous study [42], which utilized the approach of updating considered sources at every epoch based on the discriminator loss and demonstrated that limiting the number of sources to around half exhibited the best results for neural signal-based classification tasks, our subset was also restricted to half of the source participants at each epoch. Thus, participants with lower MMD measures after each epoch, calculated using conditional features derived from the feature extractor and regressor of our N2GCF, comprised the subset domain for consideration in the subsequent epoch.

### 4.7 Framework training and performance evaluation

Training and evaluation of our N2GCF were performed in a cross-subject manner, where each participant served once as the target while data from other participants were used as sources. Among the three trials performed from each source participant, the earliest recorded trial was used as training data, and the intermediate trial was used as validation data specifically for our validation loss-based model selection in an unsupervised condition (Figure 5 b). Labels from the training and validation datasets of each source participant were rescaled such that the highest label value in the training data was mapped to 1, whereas no weight shifts would be represented as a value of 0. The validation data were rescaled accordingly based on the ratio established in the training dataset. For the target participant, among the three recorded trials, the intermediate trial and the latest recorded trial were used as calibration and follow-up datasets, respectively. The highest label value from the calibration data was mapped to 1, and the follow-up data were rescaled accordingly using the same ratio derived from the calibration set. This process was done in order to match the label ranges between the training data from each source participant and the calibration data from the target participant. In cases where labeled data from the target participant’s earliest trial were used for model training, which covers the case where a model is trained in a subject-independent manner, the earliest trial was also rescaled proportionally using the ratio derived from the second trial. Overall, the data were used in chronological order, with trials recorded earlier primarily used for training the model and trials recorded later used for calibration. This was done to resemble a real-world scenario in which the data retrieved earlier are used to establish the model that would be used toward more recently acquired data.

For training our framework, the training and calibration data from the source participants and the target participant, respectively, were used in pairs to learn the parameters of the feature extractor, regressor, and domain discriminator. The training and calibration data pairs from the source participants and the target participant were also used for our MMD-based recurrent source selection and for MMD-based model selection. The calibration and follow-up datasets from the target participant were used to evaluate the framework, representing prediction error for calibration and prediction error after substantial amount of time elapsed since calibration. To quantify prediction error using the calibration and follow-up datasets, we calculated both mean absolute error (MAE) and mean squared error (MSE) between predicted and actual gait performance labels (Figure 5 c), which are the two measures widely used for regression model evaluation [61]. Both MAE and MSE were assessed to provide broader insight, with MAE serving as an intuitive error measure and MSE being more sensitive to larger errors. The samples from the calibration and follow-up datasets were composed using a time-point stride of 0.1 seconds. Note that for processes involving pairs of data between the sources and the target, each data sample from the sources was paired with a data sample from the target prior to use. These sample pairs were determined in a randomized manner and were constantly renewed at each epoch, indicating that the sample pairs would change from epoch to epoch. This approach was taken in order to match the number of samples between each source and target pair and to simplify the framework’s training process.

### 4.8 Conditions and metrics for framework model establishments

We explored the performance of our framework using two distinct conditions: unsupervised and semi-supervised. In the unsupervised condition, none of the data labels from the target participant were considered for model training. Data labels were also not used for model selection, which is the process of selecting a single model from the models established throughout training epochs. Similarly for the semi-supervised condition, none of the data labels from the target participant was used for model training, however, these labels were utilized for model selection to identify the optimally calibrated model among those established throughout the training epochs. Specifically, in the semi-supervised condition, we chose the model from the epoch that exhibited the lowest L1 loss with the calibration dataset of the target participant. With the semi-supervised condition aiming to explore our framework architecture’s ability to extract domain-invariant features independent of model selection approaches, and the unsupervised condition aiming to investigate the performance of our framework in realistic applications where the model would be calibrated without access to data labels from the target participant, we utilized both conditions to analyze the performance of our framework in greater depth.

Considering that the approach used for model selection among the models established across epochs is another important component in the unsupervised condition [62,63], we additionally utilized two different selection methods for the analysis: selecting the model with the lowest L1 loss rate using validation data from the source participants, and selecting the model based on our approach of using MMD measure between the sources’ training data features and target’s calibration data features. Specifically in terms of our MMD-based metric, the concatenated vector of conditional and marginal features were used for model selection to consider both conditional and marginal alignments between source and target data, and the model with the lowest average of the resulting MMD measures across all source participants was selected as the final model. The performance of the models obtained using our MMD-based metric was compared with those from the loss-based selection using the sources’ validation data.

### 4.9 Framework ablation study and benchmark comparisons

We further performed an ablation study to explore the effectiveness of our framework by comparing its performance over other possible design choices. We compared six different designs derived during our framework development: baseline model (BM), baseline model with recurrent source selection (BMRS), our domain adversarial training framework structure with no recurrent selection (DANS), framework with recurrent-selected subset applied only from domain discriminator to feature extractor (DARS DW), frame-work with recurrent-selected subset applied only to the regressor and feature extractor (DARS RW), and our complete N2G calibrator framework where the subset was applied to both discriminator-related and regressor-related parts (N2GCF). In baseline-based designs (BM and BMRS), the models only included the feature extractor and regressor, without domain discriminator, indicating that no adversarial training was applied to the model. All three components were utilized in other design choices, and their designs differed based on whether recurrent source selection was applied to backpropagation from the regressor, or from the domain discriminator, or both, or none.

In addition to the aforementioned designs, we adapted five domain adaptation approaches used in classification tasks within computer vision and EEG signal processing for our regression task. The key elements of these approaches from previous works can be described as follows:

- DANN [29]: The work proposed an adversarial training approach for domain adaptation, introducing a gradient reversal layer in between the feature extractor and the domain classifier to learn domain-invariant features.
- MDAN [64]: The study employs multiple domain classifiers, with each classifier designed to discriminate between its assigned source and target domain, thereby accommodating datasets involving multiple sources.
- CDAN [51]: This approach utilizes a conditional domain discriminator that aims to discriminate source and target domains based on conditional features derived from the feature extractor and the outputs from the task classifier.
- DJDAN [41]: This approach employs both a global domain discriminator and a local domain discriminator to consider both marginal and conditional distributions, using a dynamic adversarial factor that adjusts the weight between the two discriminators.
- SMDA [42]: The work uses multiple pairs of marginal and conditional discriminators, with each pair associated with a single source domain, to train the model using data from multiple sources.

Our feature extractor, domain discriminator, and regressor architecture was utilized as the backbone structure for the approaches we compared, and the output from the regressor replaced the classifier output of the original work for deriving conditional features. This adaptation was held as these approaches were originally designed for classification tasks, whereas our gait tracking task warrants a regression framework. Thus, this context should be considered when interpreting our benchmark comparison results.

### 4.10 Extended analysis

To investigate how recurrent selection occurred for our N2GCF training, we analyzed how frequent each source subject was selected across training epochs until the minimal L1 loss was achieved on the calibration data of the target, as well as how selections varied throughout the epochs. A sensitivity analysis was also conducted on our framework with varying kernel bandwidths for our MMD-based approaches to investigate the influence of MMD parameters on the calibrator’s performance. To additionally provide insight into the clinical relevance of our model, we used Kendall’s coefficient to quantify the strength of the association between our model’s predictions and our derived gait performance measures. We compared the association to that between beta power and gait performance, as beta power is a widely established biomarker for PD-related motor symptoms [10–12]. The beta power was calculated from LFPs using the same 2-second time windows, corresponding to the intervals of gait performance measurement from the force plates.

### 4.11 Settings for framework training

The methods and analysis scripts were implemented in Python, and the components of our framework were designed using Pytorch. The model was trained with the NVIDIA GeForce RTX 4090 GPU. We utilized adaptive moment estimation (ADAM) for optimization and used a batch size of 16 for the source domain. The maximum number of epoch for training was limited to 200 and model selection was held only on models established after the first 5 epochs. Adopting previous work that applied domain adversarial training [29], the learning rate was adjusted using the following formula to maintain a low error rate on source domains throughout adaptation toward the target domain:

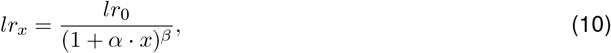

where *x* represents the proportion of elapsed batches relative to the total number of batches considering the maximum epoch limit, and *α* and *β* are constants of 10 and 0.75, respectively. The initial learning rate *lr*_0_ was set to 1e-5 in this study.

### 4.12 Statistical analysis

We utilized non-parametric tests to account for the relatively small number of result samples. For statistical comparisons regarding the error rate results retrieved from the three representative models under unsupervised and semi-supervised conditions, we used the Wilcoxon signed-rank test for pairwise comparisons between each model pairs. We conducted and reported the results using the Friedman test for multiple comparisons as a reference. The Wilcoxon signed-rank test was also used for pairwise comparisons of the Kendall correlation coefficients involving beta power and the N2G calibrator’s predictions from the two conditions. Effect size *r* values were reported to provide additional information regarding the magnitude of the effect, where values greater than 0.5 were considered large effects for our study. Bonferroni correction was applied to adjust for multiple comparisons, and the threshold for significance was defined as 0.05 after the correction.

## 5 Declarations

### 5.1 Data availability

The datasets used for the current study are not publicly available, but may be available to qualified researchers from the corresponding author upon reasonable request.

### 5.2 Code availability

The details regarding our N2G calibrator framework are presented in the Methods section of the manuscript, and the pseudocode for the related training procedures is available in our supplementary information. The codes used in the study are not publicly available, but may be available to qualified researchers from the corresponding author upon reasonable request.

## 5.3 Acknowledgements

The authors would like to thank the members of the Human Motor Control and Neuromodulation Laboratory, Dr Jaimie Henderson, and, most importantly, the participants who dedicated their time to this study. This work was supported in part by the following: NINDS UH3NS107709, NINDS UG3NS128150, NINDS UH3NS128150, NINDS R21 NS096398-02, Michael J. Fox Foundation (9605), Robert and Ruth Halperin Foundation, John A. Blume Foundation, John E Cahill Family Foundation, and Medtronic PLC who provided the devices used in this study but no additional financial support.

## 5.4 Author contributions

J.C.: Conceptualized the study, developed and analyzed deep learning models, performed data curation, wrote the original draft and revised the manuscript. H.B.S.: Conceptualized the study, revised the manuscript, and supervised the work. All authors reviewed and approved the final manuscript.

### 5.6 Corresponding author

Correspondence to Helen M. Bronte-Stewart.

---

**Supplementary Figure 1:**
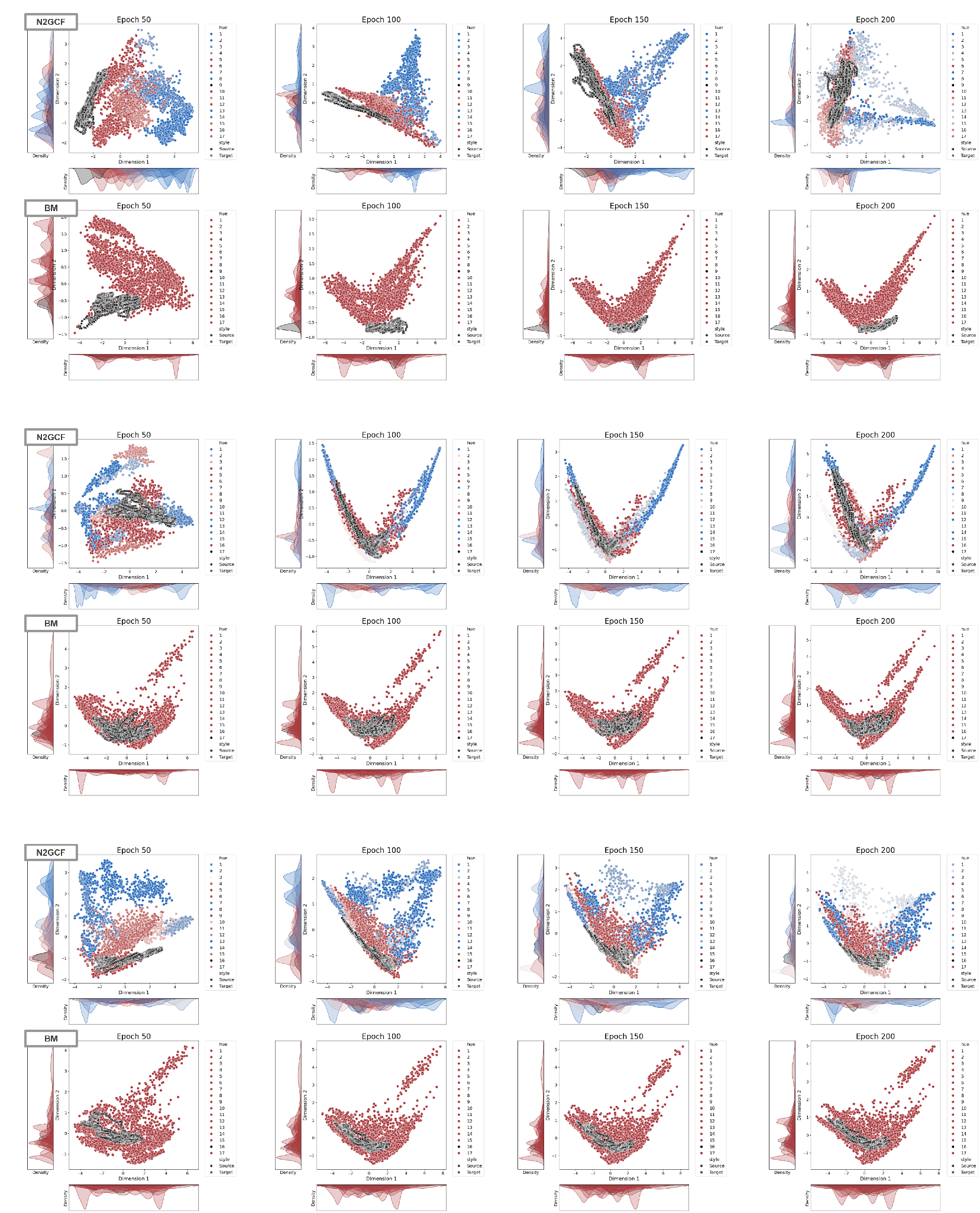
Distribution of extracted features from the sources and the appointed target across the training epochs, comparing when our adversarial training strategy was applied (N2GCF) to when the baseline model was used (BM). Each of the two rows shows instances from different participants appointed as a target user. Dimensionality reduction of features were held using principal component analysis (PCA). The color of data points represents the recurrence ratio of each source being chosen for recurrent selection until the corresponding epoch, where blue represents less frequent selection and red represents more frequent selection.

**Supplementary Figure 2:**
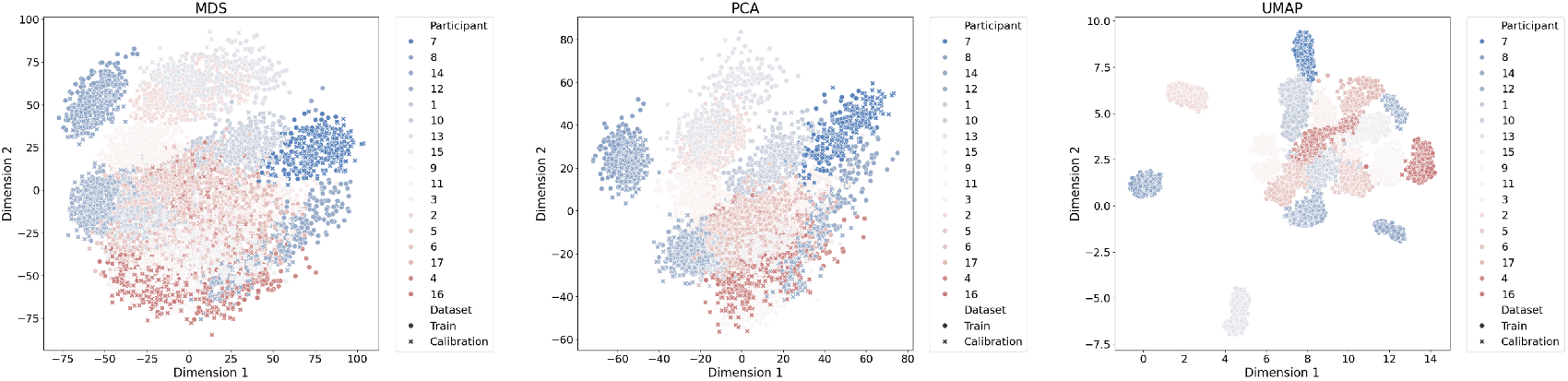
Data visualizations of neural signal data samples from all participants using the power spectral density (PSD) in the 8 and 100 Hz range. Three dimensionality reduction methods, multidimensional scaling (MDS), principal component analysis (PCA), and uniform manifold approximation and projection (UMAP), were used for dimensionality reduction. The color of data points from participants corresponds to the frequency of them being used for model training until the epoch when the minimum L1 loss was reached on the calibration data, where blue represents less frequent selection and red represents more frequent selection. The shape of the data points distinguishes whether the data points are from the training data or the calibration data.

## Algorithm 1

Training Procedure for N2G Calibration Framework

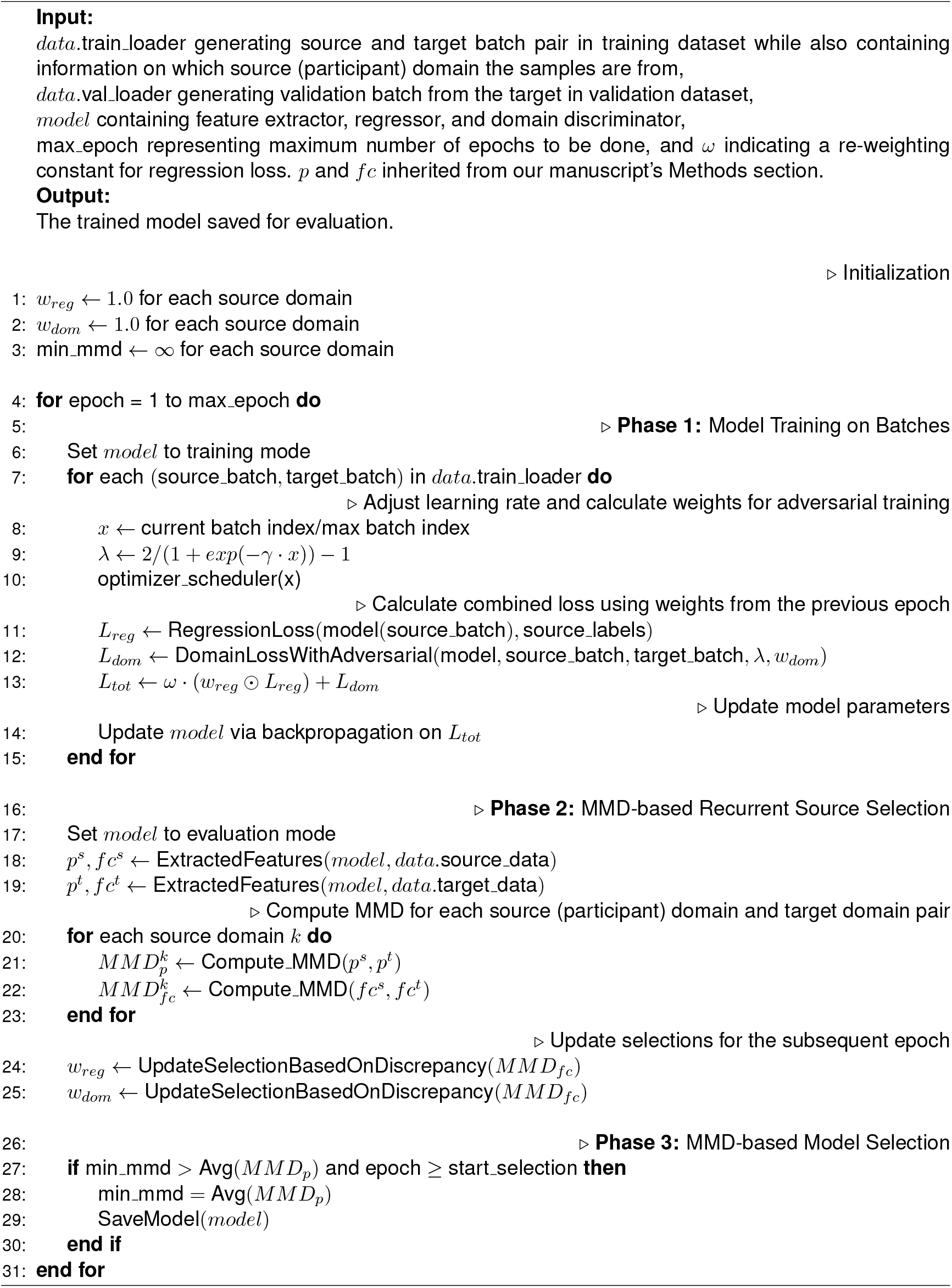

## References

[1] Priori, A., Foffani, G., Rossi, L. & Marceglia, S. Adaptive deep brain stimulation (adbs) controlled by local field potential oscillations. Experimental neurology 245, 77–86 (2013).

[2] Houston, B., Blumenfeld, Z., Quinn, E., Bronte-Stewart, H. & Chizeck, H. Long-term detection of parkinsonian tremor activity from subthalamic nucleus local field potentials. In 2015 37th Annual International Conference of the IEEE Engineering in Medicine and Biology Society (EMBC), 3427–3431 (IEEE, 2015).

[3] Opri, E. et al. Chronic embedded cortico-thalamic closed-loop deep brain stimulation for the treatment of essential tremor. Science translational medicine 12, eaay7680 (2020).

[4] Little, S. et al. Adaptive deep brain stimulation in advanced parkinson disease. Annals of neurology 74, 449–457 (2013).

[5] Beudel, M. & Brown, P. Adaptive deep brain stimulation in parkinson’s disease. Parkinsonism & related disorders 22, S123–S126 (2016).

[6] Piña-Fuentes, D. et al. Acute effects of adaptive deep brain stimulation in parkinson’s disease. Brain stimulation 13, 1507–1516 (2020).

[7] Little, S. et al. Adaptive deep brain stimulation for parkinson’s disease demonstrates reduced speech side effects compared to conventional stimulation in the acute setting. Journal of Neurology, Neurosurgery & Psychiatry 87, 1388–1389 (2016).

[8] Isaias, I. U. et al. Case report: Improvement of gait with adaptive deep brain stimulation in a patient with parkinson’s disease. Frontiers in Bioengineering and Biotechnology 12, 1428189 (2024).

[9] Molina, R. et al. Closed-loop deep brain stimulation to treat medication-refractory freezing of gait in parkinson’s disease. Frontiers in human neuroscience 15, 633655 (2021).

[10] Stanslaski, S. et al. Sensing data and methodology from the adaptive dbs algorithm for personalized therapy in parkinson’s disease (adapt-pd) clinical trial. npj Parkinson’s Disease 10, 174 (2024).

[11] Wilkins, K. B. et al. Bradykinesia and its progression are related to interhemispheric beta coherence. Annals of neurology 93, 1029–1039 (2023).

[12] Little, S. et al. Bilateral adaptive deep brain stimulation is effective in parkinson’s disease. Journal of Neurology, Neurosurgery & Psychiatry 87, 717–721 (2016).

[13] Petrucci, M. N. et al. Neural closed-loop deep brain stimulation for freezing of gait. Brain stimulation 13, 1320 (2020).

[14] Wilkins, K. B. et al. Beta burst-driven adaptive deep brain stimulation improves gait impairment and freezing of gait in parkinson’s disease. medRxiv (2024).

[15] Busch, J. L. et al. Single threshold adaptive deep brain stimulation in parkinson’s disease depends on parameter selection, movement state and controllability of subthalamic beta activity. Brain stimulation 17, 125–133 (2024).

[16] Neumann, W.-J., Gilron, R., Little, S. & Tinkhauser, G. Adaptive deep brain stimulation: from experimental evidence toward practical implementation. Movement disorders 38, 937–948 (2023).

[17] Acharyya, P. et al. Closing the loop in dbs: A data-driven approach. Parkinsonism & Related Disorders 134, 107348 (2025).

[18] Choi, J. W., Cui, C., Wilkins, K. B. & Bronte-Stewart, H. M. N2gnet tracks gait performance from subthalamic neural signals in parkinson’s disease. npj Digital Medicine 8, 7 (2025).

[19] Thenaisie, Y. et al. Principles of gait encoding in the subthalamic nucleus of people with parkinson’s disease. Science translational medicine 14, eabo1800 (2022).

[20] Canessa, A., Palmisano, C., Isaias, I. U. & Mazzoni, A. Gait-related frequency modulation of beta oscillatory activity in the subthalamic nucleus of parkinsonian patients. Brain Stimulation 13, 1743–1752 (2020).

[21] Louie, K. H. et al. Cortico-subthalamic field potentials support classification of the natural gait cycle in parkinson’s disease and reveal individualized spectral signatures. eneuro 9 (2022).

[22] He, S. et al. Closed-loop deep brain stimulation for essential tremor based on thalamic local field potentials. Movement Disorders 36, 863–873 (2021).

[23] Oliveira, A. M. et al. Machine learning for adaptive deep brain stimulation in parkinson’s disease: closing the loop. Journal of Neurology 270, 5313–5326 (2023).

[24] Dixon, T. C. et al. Movement-responsive deep brain stimulation for parkinson’s disease using a remotely optimized neural decoder. Nature Biomedical Engineering 1–15 (2025).

[25] Saha, S. & Baumert, M. Intra-and inter-subject variability in eeg-based sensorimotor brain computer interface: a review. Frontiers in computational neuroscience 13, 87 (2020).

[26] Huang, G. et al. Discrepancy between inter-and intra-subject variability in eeg-based motor imagery brain-computer interface: Evidence from multiple perspectives. Frontiers in neuroscience 17, 1122661 (2023).

[27] Arlotti, M., Rosa, M., Marceglia, S., Barbieri, S. & Priori, A. The adaptive deep brain stimulation challenge. Parkinsonism & related disorders 28, 12–17 (2016).

[28] Wang, T., Shoaran, M. & Emami, A. Towards adaptive deep brain stimulation in parkinson’s disease: Lfp-based feature analysis and classification. In 2018 IEEE international conference on acoustics, speech and signal processing (ICASSP), 2536–2540 (IEEE, 2018).

[29] Ganin, Y. et al. Domain-adversarial training of neural networks. Journal of machine learning research 17, 1–35 (2016).

[30] Nath, D. et al. A comparative study of subject-dependent and subject-independent strategies for eeg-based emotion recognition using lstm network. In Proceedings of the 2020 4th International Conference on Compute and Data Analysis, 142–147 (2020).

[31] Autthasan, P. et al. Min2net: End-to-end multi-task learning for subject-independent motor imagery eeg classification. IEEE Transactions on Biomedical Engineering 69, 2105–2118 (2021).

[32] Apicella, A. et al. Toward cross-subject and cross-session generalization in eeg-based emotion recognition: Systematic review, taxonomy, and methods. Neurocomputing 128354 (2024).

[33] Cox, T. F. & Cox, M. A. Multidimensional scaling (CRC press, 2000).

[34] Hout, M. C., Papesh, M. H. & Goldinger, S. D. Multidimensional scaling. Wiley Interdisciplinary Reviews: Cognitive Science 4, 93–103 (2013).

[35] McInnes, L., Healy, J. & Melville, J. Umap: Uniform manifold approximation and projection for dimension reduction. arXiv preprint arXiv:1802.03426 (2018).

[36] Gerster, M. et al. Beyond beta rhythms: Aperiodic broadband power reflects parkinson’s disease severity–a multicenter study. bioRxiv 2025–03 (2025).

[37] Darmani, G. et al. Long-term recording of subthalamic aperiodic activities and beta bursts in parkinson’s disease. Movement Disorders 38, 232–243 (2023).

[38] Janson, A. P., Anderson, D. N. & Butson, C. R. Activation robustness with directional leads and multi-lead configurations in deep brain stimulation. Journal of neural engineering 17, 026012 (2020).

[39] Thenaisie, Y. et al. Towards adaptive deep brain stimulation: clinical and technical notes on a novel commercial device for chronic brain sensing. Journal of neural engineering 18, 042002 (2021).

[40] Tang, X. & Zhang, X. Conditional adversarial domain adaptation neural network for motor imagery eeg decoding. Entropy 22, 96 (2020).

[41] Hong, X. et al. Dynamic joint domain adaptation network for motor imagery classification. IEEE Transactions on Neural Systems and Rehabilitation Engineering 29, 556–565 (2021).

[42] Lee, J., Choi, J. W. & Jo, S. Selective multi-source domain adaptation network for cross-subject motor imagery discrimination. IEEE Transactions on Cognitive and Developmental Systems 16, 923–934 (2023).

[43] Richard, G., Mathelin, A. d., Hébrail, G., Mougeot, M. & Vayatis, N. Unsupervised multi-source domain adaptation for regression. In Machine Learning and Knowledge Discovery in Databases: European Conference, ECML PKDD 2020, Ghent, Belgium, September 14–18, 2020, Proceedings, Part I, 395–411 (Springer, 2021).

[44] Wang, Z., Zhang, W., Li, S., Chen, X. & Wu, D. Unsupervised domain adaptation for cross-patient seizure classification. Journal of Neural Engineering 20, 066002 (2023).

[45] Fan, J. et al. Unsupervised domain adaptation by statistics alignment for deep sleep staging networks. IEEE Transactions on Neural Systems and Rehabilitation Engineering 30, 205–216 (2022).

[46] You, K., Wang, X., Long, M. & Jordan, M. Towards accurate model selection in deep unsupervised domain adaptation. In International Conference on Machine Learning, 7124–7133 (PMLR, 2019).

[47] Hu, D., Luo, R., Liang, J. & Foo, C. S. Towards reliable model selection for unsupervised domain adaptation: An empirical study and a certified baseline. Advances in Neural Information Processing Systems 37, 135883–135903 (2024).

[48] Musgrave, K., Belongie, S. & Lim, S.-N. Unsupervised domain adaptation: A reality check. arXiv preprint arXiv:2111.15672 (2021).

[49] Clark, D. L. et al. Aperiodic subthalamic activity predicts motor severity and stimulation response in parkinson disease. Parkinsonism & Related Disorders 110, 105397 (2023).

[50] Voytek, B. et al. Age-related changes in 1/f neural electrophysiological noise. Journal of neuroscience 35, 13257–13265 (2015).

[51] Long, M., Cao, Z., Wang, J. & Jordan, M. I. Conditional adversarial domain adaptation. Advances in neural information processing systems 31 (2018).

[52] Farahani, A., Voghoei, S., Rasheed, K. & Arabnia, H. R. A brief review of domain adaptation. Advances in data science and information engineering: proceedings from ICDATA 2020 and IKE 2020 877–894 (2021).

[53] Lu, N., Xiao, H., Sun, Y., Han, M. & Wang, Y. A new method for intelligent fault diagnosis of machines based on unsupervised domain adaptation. Neurocomputing 427, 96–109 (2021).

[54] Han, T., Liu, C., Yang, W. & Jiang, D. Deep transfer network with joint distribution adaptation: A new intelligent fault diagnosis framework for industry application. ISA transactions 97, 269–281 (2020).

[55] Gretton, A., Borgwardt, K., Rasch, M., Schölkopf, B. & Smola, A. A kernel method for the two-sample-problem. Advances in neural information processing systems 19 (2006).

[56] Gretton, A., Borgwardt, K. M., Rasch, M. J., Schölkopf, B. & Smola, A. A kernel two-sample test. The Journal of Machine Learning Research 13, 723–773 (2012).

[57] Zhuang, F. et al. A comprehensive survey on transfer learning. Proceedings of the IEEE 109, 43–76 (2020).

[58] Long, M., Cao, Y., Wang, J. & Jordan, M. Learning transferable features with deep adaptation networks. In International conference on machine learning, 97–105 (PMLR, 2015).

[59] Mekhazni, D., Bhuiyan, A., Ekladious, G. & Granger, E. Unsupervised domain adaptation in the dissimilarity space for person re-identification. In Computer Vision–ECCV 2020: 16th European Conference, Glasgow, UK, August 23–28, 2020, Proceedings, Part XXVII 16, 159–174 (Springer, 2020).

[60] Tolstikhin, I. O., Sriperumbudur, B. K. & Schö lkopf, B. Minimax estimation of maximum mean discrepancy with radial kernels. Advances in Neural Information Processing Systems 29 (2016).

[61] Merk, T. et al. Machine learning based brain signal decoding for intelligent adaptive deep brain stimulation. Experimental Neurology 351, 113993 (2022).

[62] Fang, Y., Yap, P.-T., Lin, W., Zhu, H. & Liu, M. Source-free unsupervised domain adaptation: A survey. Neural Networks 106230 (2024).

[63] Wilson, G. & Cook, D. J. A survey of unsupervised deep domain adaptation. ACM Transactions on Intelligent Systems and Technology (TIST) 11, 1–46 (2020).

[64] Zhao, H. et al. Adversarial multiple source domain adaptation. Advances in neural information processing systems 31 (2018).

